# Determinants of HIV-1 late presentation in a cohort of Portuguese HIV-1 patients

**DOI:** 10.1101/2020.06.17.20133785

**Authors:** Ana Cláudia Miranda, Mafalda Miranda, Marta Pingarilho, Victor Pimentel, João Torres, Susana Peres, Teresa Baptista Alberto, Perpetua Gomes, Ana Abecasis, Kamal Mansinho

**Affiliations:** Serviço de Doenças Infeciosas, Centro Hospitalar de Lisboa Ocidental, Hospital de Egas Moniz, Lisboa, Portugal; Global Health and Tropical Medicine (GHTM), Instituto de Higiene e Medicina Tropical/Universidade Nova de Lisboa (IHMT/UNL), Lisboa, Portugal; Laboratório de Biologia Molecular (LMCBM, SPC, CHLO-HEM), Lisboa, Portugal; Centro de Investigação Interdisciplinar Egas Moniz (CiiEM), Instituto Superior de Ciências da Saúde Egas Moniz, Caparica, Portugal

**Keywords:** HIV-1 infection, Late presentation, Late presentation with advanced disease

## Abstract

**Background:** Undiagnosed HIV-1 patients still account for 25% of worldwide HIV patients. Studying late presenters for HIV care may help to identify characteristics of such patients.

**Objective:** The present study aims to identify factors associated with late presentation (LP) and late presentation with advanced disease (LPAD) based on a population of patients followed in a Portuguese hospital between 1984 and 2017.

**Methods:** Sociodemographic and clinical data from infected patients with HIV-1 aged 18 years and older, followed in Egas Moniz Hospital, in Portugal were collected.

**Results:** Of the 907 patients included in this study, 68.7% were males and the median age was 37 years (IQR 30-47). 459 patients (50.6%) were LP and, of these, 284 patients (61.9%) were LPAD. The LP population mostly originated from Portugal and Sub-Saharan Africa (64.4% and 28.8%; *p*=0.004) and the HIV exposure category mainly heterosexuals and MSM (57.0% and 24.9%; *p*<0.001). The stage of disease and viral load at diagnosis were significantly associated with both LP and LPAD (*p*<0.001). Factors associated with LP in the logistic regression included age at diagnosis lower than 30y (aOR 0.34; 0.17-0.68; *p*=0.002) and origin from Sub-Saharan Africa (aOR 2.24; 1.44-3.50; *p*<0.001).

**Conclusion:** Late presentation is a major obstacle to halt the HIV epidemic. In this population, the majority of newly diagnosed HIV-infected individuals were late presenters. Our results characterize vulnerable populations that should be frequently tested for HIV.

## Introduction

Human Immunodeficiency Virus (HIV) continues to be one of the main public health issues. In 2018 there was 1.7 million people newly infected worldwide and 973 new cases were reported in Portugal [1, 2]. Early diagnosis is vital to achieve the objectives proposed by the WHO: the 95-95-95 target to end the pandemic by 2030: diagnosing 95% of people living with HIV, 95% of diagnosed on treatment, and 95% of people on treatment viral suppressed [3]. However, people living with HIV who do not know their status, account for 25% of the total infected people worldwide (9.4 million people) [4]. In the European Union it is estimated that Late Presenters (LP) represent around 49-54% of cases and Late Presenters with Advanced Disease (LPAD) are around 33-42% of HIV cases [5]. According to the last Portuguese report, in 2018, LP cases accounted for 55.8% and LPAD cases accounted for 34.3% of HIV infection [2]. Importantly, the proportion of LP cases among newly diagnosed is increasing, indicating that we are leaving some older cases of undiagnosed patients behind.

According to the European Late Presenter Consensus working group, Late presenters (LP) were defined as presenting a TCD4^+^ count lower than 350 cells/mm^3^ or an AIDS defining event, regardless of TCD4^+^ cell count [6]. A subgroup of late presenters, called late presenters with advanced disease (LPAD) are characterized by presenting a TCD4^+^ count lower than 200 cells/mm^3^ or an AIDS-defining event, regardless of TCD4^+^ cell count. This latest subgroup particularly is at greater risk of severe disease and death [7,8]. The current guidelines for LP and LPAD patients, suggest that they are unable to fully benefit from antiretroviral therapy (ART), leading to poorer outcomes in treatment. This late entry to care can not only have an impact in individual’s morbidity and mortality, but also can increase the risk of onward transmission due to unawareness of their HIV status, with an impact on the control of the pandemic [9–11].

The present study has the objective of identifying determinants of late presentation (LP) and late presentation with advanced disease (LPAD). To do this, we analyzed a population of patients followed up in a Portuguese hospital, diagnosed between 1984 and 2017.

## Methods

### Study Group

Clinical and socio-demographic information from 907 HIV-1 infected patients was collected during routine clinical care of patients followed in Hospital Egas Moniz, which is part of Centro Hospitalar de Lisboa Ocidental (CHLO), Lisbon, Portugal, between 1984 and 2017. Patients were aged 18 years and over.

### Statistical analysis

Descriptive analysis was conducted. The proportion and median (interquartile range, IQR) of LP, NLP and LP-AD was calculated for every categorical and continuous variable, respectively. Our interest variables were compared with the categorical variables with Chi-square test, and continuous variables with t-test for independent samples.

To study the association between our dependent variable and the independent variables, logistic regression models were calculated. We first presented the logistic regression with the unadjusted odds ratios (uOR) and confidence intervals at 95% (95%CI), variables with a p-value <0.05 were considered to enter the model. We calculated the logistic regression model with the stepwise mode, which lead to the construction of the final model. The final model for LP vs NLP was adjusted for gender, this variable was forced into the model regardless of its significance and the reference class were women, and the final model for LP vs LP-AD was adjusted for gender and age at diagnosis. Results were considered statistically significant when *p*<0.05. The odds ratio and 95% confidence intervals were calculated for the variables of the final model. Data was analyzed using SPSS for Windows (Version 23.0).

### Ethics

The protocol was in accordance with the declaration of Helsinki and approved by the Ethical Committee of Centro Hospitalar de Lisboa Ocidental (108/CES-2014). This database contains anonymized patients’ information, including demographic and clinical data from patients followed in Hospital Egas Moniz between 1984-2017.

## Results

Among 907 HIV-1 infected patients included in the analysis the median age was 37 (IQR: 30.0-47.0) years and 68.7% were males (Table 1). 459 patients (50.6%) were LP and, of these, 284 patients (61.9%) were LPAD. Heterosexuals and Portuguese originated patients presented a higher proportion in this study population, 52.3% and 67.7% respectively. CD4 count at diagnosis and viral load at diagnosis (log_10_) presented a median of 342 cells/mm^3^ (IQR 155-554) and 4.8 copies/mL (IQR 4.2-5.4), respectively. Patients at stage A of HIV infection accounted for 66.8% of the population of patients, when compared to stages B and C.

**Table 1.** Demographic and clinical patient characteristics

| Patient Characteristics | Total | Late Presenters | Non-Late Presenters | p-value | Late Presenters with advanced disease | p-value |
| --- | --- | --- | --- | --- | --- | --- |
| **Gender, n (%)** | 907 (100%) | 459 (50.6%) | 448 (49.4%) | 0.499 | 284 (61.9%) | 0.403<sup>1</sup> |
| Female | 284 (31.3%) | 139 (30.3%) | 145 (32.4%) |  | 82 (28.9%) |  |
| Male | 623 (68.7%) | 320 (69.7%) | 303 (67.6%) |  | 202 (71.1%) |  |
| **Median age at diagnosis in years IQR, n (%)** | 806 (100%) | 430 (53.3%) | 376 (46.7%) | <0.001 | 264 (61.4%) | 0.049<sup>2</sup> |
|  | 37.0 (30.0-47.0) | 39.5 (32.0-50.0) | 33.5 (28.0-43.0) |  | 41.0 (32.25-51.0) |  |
| <30 | 230 (28.5%) | 89 (20.7%) | 141 (37.5%) | <0.001 | 46 (17.4%) | 0.049<sup>1</sup> |
| 31-55 | 488 (60.5%) | 276 (64.2%) | 212 (56.4%) |  | 172 (65.2%) |  |
| >56 | 88 (10.9%) | 65 (15.1%) | 23 (6.1%) |  | 46 (17.4%) |  |
| **Type of transmission, n (%)** | 894 (100%) | 453 (50.7%) | 441 (49.3%) | 0.001 | 278 (61.4%) | 0.496<sup>1</sup> |
| Heterosexual | 468 (52.3%) | 258 (57.0%) | 210 (47.6%) |  | 165 (59.4%) |  |
| MSM | 275 (30.8%) | 113 (24.9%) | 162 (36.7%) |  | 66 (23.7%) |  |
| **IDU** | 141 (15.8%) | 75 (16.6%) | 66 (15.0%) |  | 44 (15.8%) |  |
| **Other** | 10 (1.1%) | 7 (1.5%) | 3 (0.7%) |  | 3 (1.1%) |  |
| **Region of origin, n (%)** | 899 (100%) | 455 (50.6%) | 444 (49.4%) | **0.004** | 282 (62.0%) | 0.893<sup>1</sup> |
| **Portugal** | 609 (67.7%) | 293 (64.4%) | 316 (71.2%) |  | 181 (64.2%) |  |
| **Sub-Saharan Africa** | 215 (23.9%) | 131 (28.8%) | 84 (18.9%) |  | 80 (28.4%) |  |
| **Brasil** | 54 (6.0%) | 23 (5.1%) | 31 (7.0%) |  | 16 (5.7%) |  |
| **Other** | 21 (2.3%) | 8 (1.8%) | 13 (2.9%) |  | 5 (1.8%) |  |
| **Stage of Infection at diagnosis, n (%)** | 895 (100%) | 453 (50.6%) | 442 (49.4 %) | **<0.001** | 276 (61.3%) | **<0.001<sup>1</sup>** |
| **A** | 598 (66.8%) | 220 (48.6%) | 378 (85.5%) |  | 94 (34.1%) |  |
| **B** | 131 (14.6%) | 84 (18.5%) | 47 (10.6%) |  | 54 (19.6%) |  |
| **C** | 166 (18.5%) | 149 (32.9%) | 17 (3.8%) |  | 128 (46.4%) |  |
| **Median CD4 count at diagnosis (cells/mL) IQR, n (%)** | 907 (100%) | 459 (50.6%) | 448 (49.4%) |  | 284 (61.9%) |  |
|  | 342.0 (155.0-554.0) | 158.0 (59.0-250.0) | 555.5 (444.0-712.0) |  | 83.5 (33.0-144.0) |  |
| Viral Load at diagnosis (log<sub>10</sub> copies/mL) IQR, n (%) | 785 (100%) | 396 (50.4%) | 389 (49.6%) | <0.001 | 241 (60.9%) | <0.001<sup>2</sup> |
|  | 4.8 (4.2-5.4) | 5.1 (4.7-5.6) | 4.4 (3.8-4.9) |  | 5.3 (4.9-5.7) |  |
| >4.0 | 146 (18.6%) | 27 (6.8%) | 119 (30.6%) | <0.001 | 11 (4.6%) | <0.001<sup>1</sup> |
| 4.1-5.0 | 322 (41.0%) | 139 (35.1%) | 183 (47.0%) |  | 61 (25.3%) |  |
| <5.1 | 317 (40.4%) | 230 (58.1%) | 87 (22.4%) |  | 169 (70.1%) |  |

Other in mode of transmission include transfusions; Other in region of origin include European, Latin and North American Countries; p-values retrieved with t-test and chi-square test; MSM- Men have sex with men; IDU- Injection drug users

Characteristics of patients stratified according to time of presentation are presented in Table 1. In this sample, males accounted for the bigger proportion of LP (69.7%); 71.1% of those being LPAD. No statistical differences according to gender were found for LP and LPAD populations. The median age for LP was 39.5 (IQR 32-50; *p*<0.001) and the age group between 31-55yo (*p*<0.001) represented 64.2% of LP, significantly higher when compared with the other age groups (≤30 and ≥56; *p*<0.001). The median age for LPAD, 41 (IQR 32.25-51; *p*=0.049), was higher than LP. The median CD4 count for LP was 158 cell/mm^3^ (IQR 59-250) and for LPAD was 83.5 cells/mm^3^ (IQR 33-144). LP population was mainly from Portugal and Sub-Saharan Africa (64.4% vs 28.8%) and the HIV exposure category were mainly heterosexuals and MSM (57.0% vs 24.9%). In the univariate analysis, both region of origin and HIV exposure category were associated with LP (*p*<0.001), but not associated with LPAD. Clinical characteristics, as stage of disease and viral load at diagnosis were associated with both LP and LPAD (*p*<0.001), for LP stage A had the higher proportion 48.6%, but for LPAD the stage C had higher percentage, 46.4%.

In the unadjusted model (Table 2), for LP vs non-LP, no significant differences were found between gender. In the HIV exposure category, significant differences were found for MSM compared with heterosexuals, with a higher proportion of heterosexuals among LP. Furthermore, significantly more immigrants from Sub-Saharan Africa were LP when compared to native Portuguese. In LP vs LPAD unadjusted model significant differences were found in stage of infection, youngest age group compared to the older one (LPAD are older) and viral loads higher than 5.1 compared to viral loads lower than 4.0, as consistent with evolution of infection. However, no significant differences were found between gender, HIV exposure category and region of origin.

**Table 2.** Univariate and multivariate logistic regression analysis of factors associated with late presentation and late presentation with advanced disease

|  |  | LP vs NLP |  |  |  | LP vs LPAD |  |  |  |
| --- | --- | --- | --- | --- | --- | --- | --- | --- | --- |
|  |  | Unadjusted |  | Final Model |  | Unadjusted |  | Final Model |  |
|  |  | OR (95%CI) | p-value | aOR (95%CI) | p-value | OR (95%CI) | p-value | aOR (95%CI) | p-value |
| Sex | Female | Ref |  | Ref |  | Ref |  | Ref |  |
|  | Male | 1.10 (0.83-1.46) | 0.499 | 1.07 (0.73-1.58) | 0.732 | 1.19 (0.79-1.79) | 0.403 | 0.91 (0.54-1.51) | 0.706 |
| Age at diagnosis |  | 1.04 (1.03-1.05) | **<0.001** |  |  | 1.02 (1.00-1.03) | **0.048** |  |  |
| Age groups | <30 | 0.22 (0.13-0.39) | **<0.001** | 0.34 (0.17-0.68) | **0.002** | 0.44 (0.22-0.87) | **0.018** | 0.60 (0.27-1.35) | 0.216 |
|  | 31-55 | 0.46 (0.28-0.77) | **0.003** | 0.54 (0.19-1.03) | 0.060 | 0.68 (0.38-1.23) | 0.203 | 0.72 (0.36-1.43) | 0.347 |
|  | >56 | Ref |  | Ref |  | Ref |  | Ref |  |
| Type of transmission | Heterosexual | Ref |  | Ref |  | Ref |  | Ref |  |
|  | MSM | 0.57 (0.42-0.77) | **<0.001** |  |  | 0.79 (0.50-1.24) | 0.311 |  |  |
|  | IDU | 0.93 (0.63-1.35) | 0.686 |  |  | 0.80 (0.47-1.35) | 0.405 |  |  |
|  | Other | 1.90 (0.49-7.44) | 0.357 |  |  | 0.42 (0.09-1.93) | 0.266 |  |  |
| Region of origin | Portugal | Ref |  | Ref |  | Ref |  |  |  |
|  | Sub-Saharan Africa | 1.68 (1.23-2.31) | **0.001** | 2.24 (1.44-3.50) | **<0.001** | 0.97 (0.64-1.48) | 0.890 |  |  |
|  | Brasil | 0.80 (0.46-1.40) | 0.437 | 0.68 (0.31-1.51) | 0.349 | 1.41 (0.56-3.55) | 0.460 |  |  |
|  | Other | 0.66 (0.27-1.62) | 0.369 | 0.47 (0.15-1.43) | 0.181 | 1.03 (0.24-4.40) | 0.967 |  |  |
| Stage of infection at diagnosis | A | Ref |  | Ref |  | Ref |  | Ref |  |
|  | B | 3.07 (2.07-4.55) | **<0.001** | 2.95 (1.79-4.86) | **<0.001** | 2.41 (1.43-4.06) | **0.001** | 2.42 (1.35-4.35) | **0.003** |
|  | C | 15.06 (8.88-25.55) | **<0.001** | 10.16 (5.38-19.19) | **<0.001** | 9.53 (5.44-16.71) | **<0.001** | 7.01 (3.79-12.98) | **<0.001** |
| Viral load at diagnosis |  | 2.71 (2.23-3.30) | **<0.001** |  |  | 2.08 (1.57-2.75) | **<0.001** |  |  |

| Viral load groups | <4.0 | Ref |  | Ref |  | Ref |  | Ref |  |
| --- | --- | --- | --- | --- | --- | --- | --- | --- | --- |
|  | 4.1-5.0 | 3.35 (2.09-5.37) | <0.001 | 3.40 (2.00-5.79) | <0.001 | 1.14 (0.49-2.63) | 0.763 | 1.14 (0.45-2.90) | 0.788 |
|  | >5.1 | 11.65 (7.17-18.93) | <0.001 | 7.01 (4.03-12.20) | <0.001 | 4.03 (1.77-9.16) | 0.001 | 3.10 (1.24-7.77) | 0.016 |

Other in mode of transmission include transfusions; Other in region of origin include European, Latin and North American Countries; p-values retrieved with t-test and chi-square test; MSM- Men have sex with men; IDU- Injection drug users; Ref- Reference Category

In the adjusted model, age at diagnosis is one of the factors associated with late presentation (Table 2), patients with less than 30yo had lower probability of being LP than patients with >56yo (aOR 0.34; 0.17-0.68; *p*=0.002). Patients from Sub-Saharan Africa had 2.24 more probability of presenting late than those from Portugal (aOR 2.24; 1.44-3.50; *p*<0.001) and patients presenting stage B or C had higher probability of being LP than those in stage A (aOR 2.95; 1.79-4.86 and aOR 10.16; 5.38-19.19); *p*<0.001 and *p*<0.001, respectively). The last variable associated with late presentation was viral load at diagnosis, patients with a viral load between 4.1-5.0 and >5.1 had higher probability than those with a viral load of <4.0 (aOR 3.40; 2.00-5.79 and aOR 7.01; 4.03-12.20; *p*<0.001 and *p*<0.001, respectively).

In the LPAD model (Table 2) the factors associated with late presentation with advanced disease included stage of infection at diagnosis - patients presenting stage B or C had higher probability than those in stage A (aOR 2.42; 1.35-4.35 and aOR 7.01; 3.79-12.98); *p*=0.003 and *p*<0.001, respectively) and viral load at diagnosis - patients with a viral load between >5.1 had higher probability of being LPAD than those with a viral load of <4.0 (aOR 3.10; 1.24-7.77; *p*=0.016).

## Discussion

This study had the goal of understanding the determinants of late presentation for HIV-1 infection.

In our population of patients, late presenters represented 50.6% of the patients. Of these, 61.9% were late presenters with advanced disease, which is consistent with the last national report (2017) [2]. Our results are in accordance with overall European data (2017), in which the late presenters account for 49% of the HIV cases, and 28% were late presenters with advanced disease [12].

The proportion of late presenters were higher in male gender, patients with heterosexual transmission, immigrants originated from Sub-Saharan Africa and patients aged between 31 and 55 years old. These results are consistent with other studies [8,11,13]. Patients originated from Sub-Saharan Africa represent 23.9% of the study population, of those 28.8% were LP, which is substantially lower when compared to a similar Belgian study, where the proportion of patients from Sub-Saharan African represented 54.3% of the total. [14].

According to our results the stage A of infection had higher proportions for the LP population, while in the LPAD population the higher proportion was for the stage C. This is expected and can be considered as a partial validation for our assignment of LP and LPAD to patients groups [15].

The results from previous studies showed statistically significant correlation between late presentation and HIV exposure category [6,16,17]. While we did not find this significant correlation in our logistic regression analysis, we did find it in the univariate analysis. However, recent changes in the proportion of HIV exposure categories among new diagnoses could confound such analysis [18,19].

The main goal of this study was to identify factors associated with late presentation and late presentation with advanced disease. Those factors included age at diagnosis, region of origin, stage of infection at diagnosis and log10 of the viral load at diagnosis and were in concordance with published studies [10,20] [11].

## Conclusion

Even though Portugal has achieved the 90-90-90 objectives, the proportion of late presenters is still very high, indicating that vulnerable populations are being left behind in screening protocols. Late presentation is a high impact issue at individual, economic and social level. Our study highlighted the main factors associated with that condition. Targeted prevention and screening programs should be directed to these population.

## Data Availability

No data available

## Funding

This study was financed by FCT through the following projects: MigrantsHIV PTDC/DTP-EPI/7066/2014, BESTHOPE HIVERA: Harmonizing Integrating Vitalizing European Research on HIV/Aids, grant 249697 and GHTM-UID/Multi/04413/2013 and Gilead Génese HIVLatePresenters.

## Notes

### Competing Interest Statement

The authors have declared no competing interest.

### Author Declarations

The protocol was in accordance with the declaration of Helsinki and approved by the Ethical Committee of Centro Hospitalar de Lisboa Ocidental (108/CES-2014). This database contains anonymized patients information, including demographic and clinical data from patients followed in Hospital Egas Moniz between 1984-2017.

